# Clinical Relevance of Computationally Derived Attributes of Arteries and Arterioles in focal segmental glomerulosclerosis and minimal change disease

**DOI:** 10.1101/2025.10.08.25336276

**Authors:** Jin Zhou, Dawit Demeke, Xiang Li, Timothy Dinh, Christopher O’Connor, Jane Liu, Jarcy Zee, Takaya Ozeki, Yijiang Chen, Andrew R. Janowczyk, Lawrence Holzman, Laura H. Mariani, Markus Bitzer, Laura Barisoni, Jeffrey B. Hodgin, Kyle J. Lafata

## Abstract

**Background:** The current semi-qualitative methods used to score sclerosis and hyalinosis in arteries and arterioles in clinical practice are limited in standardization and reproducibility. We developed a computational pipeline designed to accurately and consistently quantify prognostic arterial and arteriolar characteristics in digital kidney biopsies of patients with focal segmental glomerulosclerosis (FSGS) and minimal change disease (MCD) through segmentation and pathomic feature extraction.

**Methods:** We utilized one trichrome-stained WSI from 225 participants in the NEPTUNE/CureGN studies, comprising 127 cases of focal segmental glomerulosclerosis (FSGS) and 98 cases of minimal change disease (MCD). We developed, validated, and quality-controlled deep learning models to segment muscular vessels and their internal compartments (lumen, intima, media, and hyalinosis), including (i) arcuate arteries, (ii) interlobular arteries, and (iii) arterioles with two muscle layers. Arterioles, interlobular, and arcuate arteries were visually scored for sclerosis and hyalinosis on a scale of 0 to 3. Area- and thickness-based pathomic feature extraction was performed on each compartment (lumen, intima, media, and hyalinosis) through radial sampling and ray casting. A correlation study was performed between pathomic and visual semiquantitative visual scores, and the association of both visual scores and pathomic features with disease progression (40% eGFR decline or renal failure) was assessed. Summary statistics (maximum, median, and 75th percentile) were computed for each WSI and analyzed using LASSO-regularized Cox proportional hazards models, adjusted for clinical and demographic factors.

**Results:** A total of 1,499 arterioles, 686 interlobular arteries, and 131 arcuate arteries were segmented. Statistically significant correlations were found between pathologists visual scores and the average intima-media thickness ratio (Spearman ρ = 0.27, p < 0.001 for arterioles; ρ = 0.69, p < 0.001 for interlobular arteries; and ρ = 0.80, p < 0.001 for arcuate arteries) and arteriolar hyalinosis (ρ = 0.46, p < 0.001). Incorporating pathomic features from trichrome-stained WSIs improved the prediction of disease progression, enhancing the concordance index from 0.70 to 0.75 in arterioles and from 0.69 to 0.74 in arcuate arteries, compared to using demographics and clinical characteristics alone.

**Conclusion:** Our computational approach offers a novel and reliable method for segmenting and analyzing the pathomic features of sclerosis and hylalinosis in arteries and arterioles. This technique has demonstrated potential as a valuable tool for enhancing the clinical assessment performed by pathologists.

**Key Points:**

1. A computational pipeline was developed and validated to segment arteries and arterioles and to quantify lumen, intima, media, and hyalinosis in kidney biopsies from patients with FSGS and MCD.
2. Pathomic features, such as intima-media thickness ratio and hyalinosis area, significantly correlated with pathologists’ semi-quantitative sclerosis and hyalinosis scores.
3. Integrating pathomic features into clinical models improved disease progression prediction accuracy

## Introduction

Recent advancements in digital pathology, combined with access to large digitized kidney biopsies and extensive clinical datasets, provide unprecedented opportunities to precisely and consistently quantify histologic parameters and assess their clinical relevance in research, trials, and routine practice.^1–8^ Computer vision can be applied to transform whole slide images (WSI) into actionable data, allowing for the automatic extraction of information beyond visual representation. This information can be used to characterize tissues better, enhancing pathologists’ capabilities in diagnosing, prognosticating, and predicting disease outcomes.^3,9–15^ In the kidney, computational approaches have increasingly been applied to structurally characterize and quantify functional tissue units such as glomeruli, tubules, arteries, peritubular capillaries, etc., through deep learning segmentation techniques and pathomic feature extraction.^14,16,17^ While recent studies have focused on glomeruli^18,19^, tubules^17,20^, peritubular capillaries^16^, and regions of interstitial fibrosis and tubular atrophy^21^, pathomics-derived characterization of arteries/arterioles has not yet been fully explored.^22^

In clinical practice, current approaches for the assessment of arteries/arterioles in kidney biopsies is based on the visual semiquantitative visual scoring of arteriosclerosis and hyalinosis.^23–25^ However, this approach may be influenced by inherent cognitive and visual biases,^26^ with the precision and accuracy of these observations limited by high inter- and intra-observer variability.^27–30^ There is also a lack of universal standardization, for example, some pathologists use a gestalt of the severity of arteriosclerosis across all arteries/arterioles present in the biopsy, while others base their score on the most severe lesion.^28^ Although several techniques have been proposed for the automatic characterization of arteriosclerosis and hyalinosis, they often simplify the underlying biological complexities or have not been fully explored for their generalized prognostic value.^22,25^

This study aims to develop a computational approach to quantify prognostic arterial/arteriolar characteristics in digital kidney biopsies of patients with focal segmental glomerulosclerosis (FSGS) and minimal change disease (MCD) through segmentation and pathomic feature extraction from arteries/arterioles subcompartments (lumen, intima, media).

## Methods

### Study Sample Selection

This study leveraged the infrastructure of the Nephrotic Syndrome Study Network (NEPTUNE) and Cure Glomerulonephropathies (CureGN) consortia. NEPTUNE is a multisite observational cohort study of children and adults with proteinuria, enrolled at the time of their first clinically indicated kidney biopsy.^31^ A fraction of NEPTUNE participants are also enrolled in CureGN. Inclusion criteria: (i) diagnosis of focal segmental glomerulosclerosis (FSGS) or minimal change disease (MCD) including classic MCD and MCD-like (presence of global sclerosis and/or partial foot process effacement)^32^, (ii) a digitized renal biopsy in the NEPTUNE/CureGN digital pathology repository, (iii) a good quality trichrome-stained WSI, (iv) presence of arteries in the selected WSIs, (v) available demographic data: age, sex, and self-reported race or race reported by parents for children, (vi) available clinical data: estimated glomerular filtration rate (eGFR) and urine protein creatinine ratio (UPCR) at the time of biopsy, (vii) follow-up data on changes in eGFR and kidney replacement therapy, and (viii) biopsy-level semiquantitative visual scores (0-3) for arteriosclerosis and hyalinosis, extracted from the NEPTUNE/CureGN data repository (NEPTUNE/CureGN core scoring).

### Arterial/Arteriolar Segmentation, Categorization and Scoring

#### Segmentation

For each WSI, all muscular vessels, along with their intra-vascular compartments (media, intima, lumen), were segmented using previously validated deep learning models.^22,33^ The results were then imported into QuPath,^34^ where pathologists quality-controlled the segmentations by manually adjusting the boundaries of each segmented area. Additionally, areas of hyalinosis were annotated by the pathologists during the quality control process.

#### Categorization & Scoring

For manual categorization and scoring for arteriosclerosis and arteriolar hyalinosis severity we used an online platform, Labelbox^35^. All previously segmented muscular vessels were categorized on their size and number of smooth muscle layers into arcuate and interlobular arteries, arterioles, and muscular venules (Figure 2). Specifically, for arterioles, only those with two muscle layers were analyzed, while arterioles with a single muscle layer and muscular venules were excluded from the study. Additionally, any artery/arteriole displaying an absence of lumen or artifacts was also excluded from the analysis. Arcuate arteries were identified as large vessels at the corticomedullar junction and generally surrounded by ample adventia. Interlobular arteries were characterized by >2 layers of smooth muscle cells, of intermediate size between arterioles and arcuate arteries, and clearly within the cortex.

**Figure 1.**
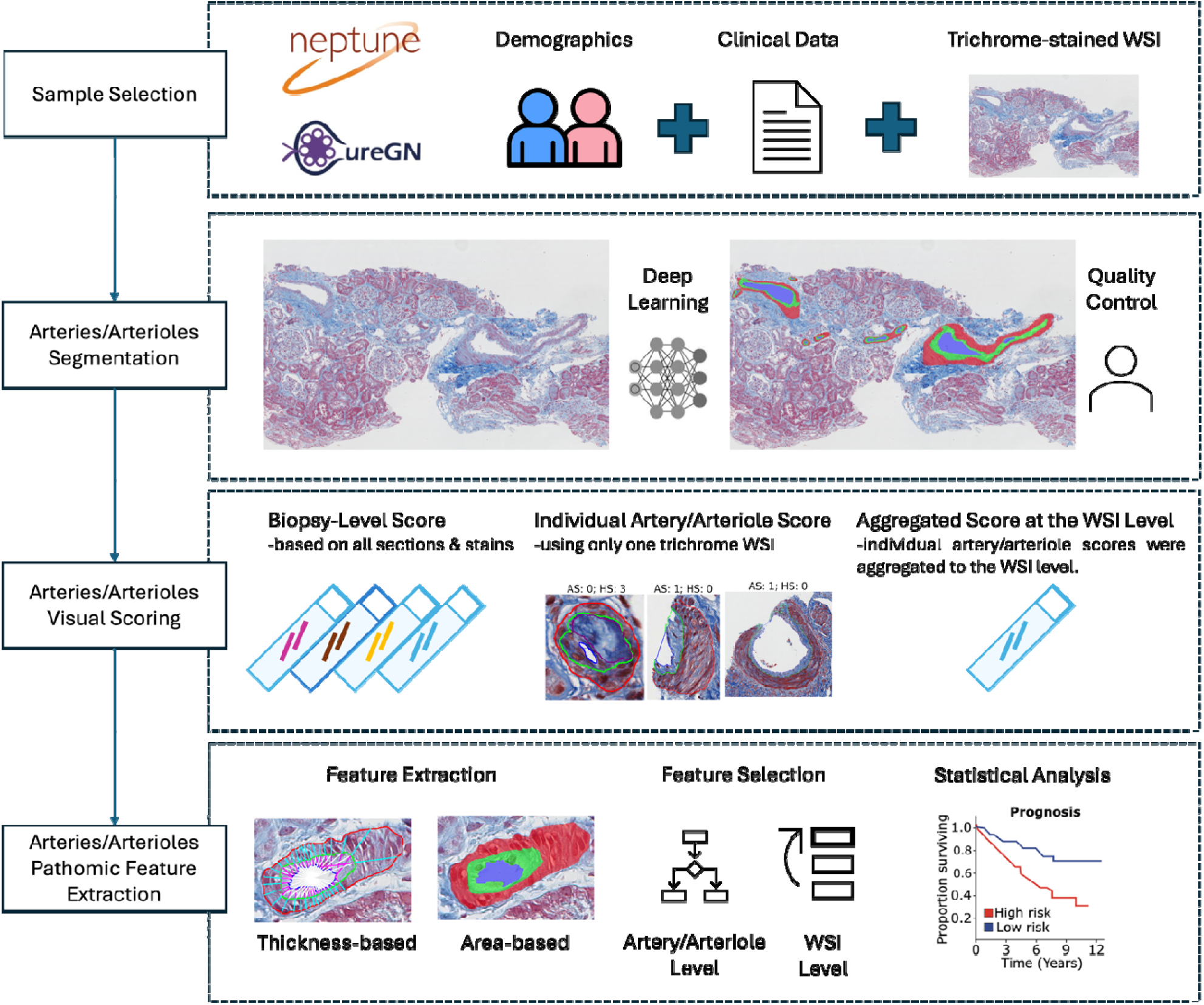
Study design overview for arterial/arteriolar characterization. First Panel: Demographics, clinical data and trichrome-stained whole slide images from the NEPTUNE and CureGN dataset were used. Second Panel: Deep learning models were developed to segment muscular vessels and intra-arterial/arteriolar compartments, followed by manual annotation of hyalinosis and quality control of boundaries. Third Panel: Severity of arteriosclerosis and hyalinosis was assessed using a semiquantitative visual score as follow: (1) visually, at the biopsy-level (all sections and stains); (2) visually, at the individual artery/arteriole (one trichrome WSI per biopsy); and (3) aggregated score at the trichrome WSI level. Fourth Panel: Computational quantification of arterial/arteriolar intima and media thickness and area measurements, feature selection (artery/arteriole and WSI levels), and assessment of the clinical relevance.

**Figure 2.**
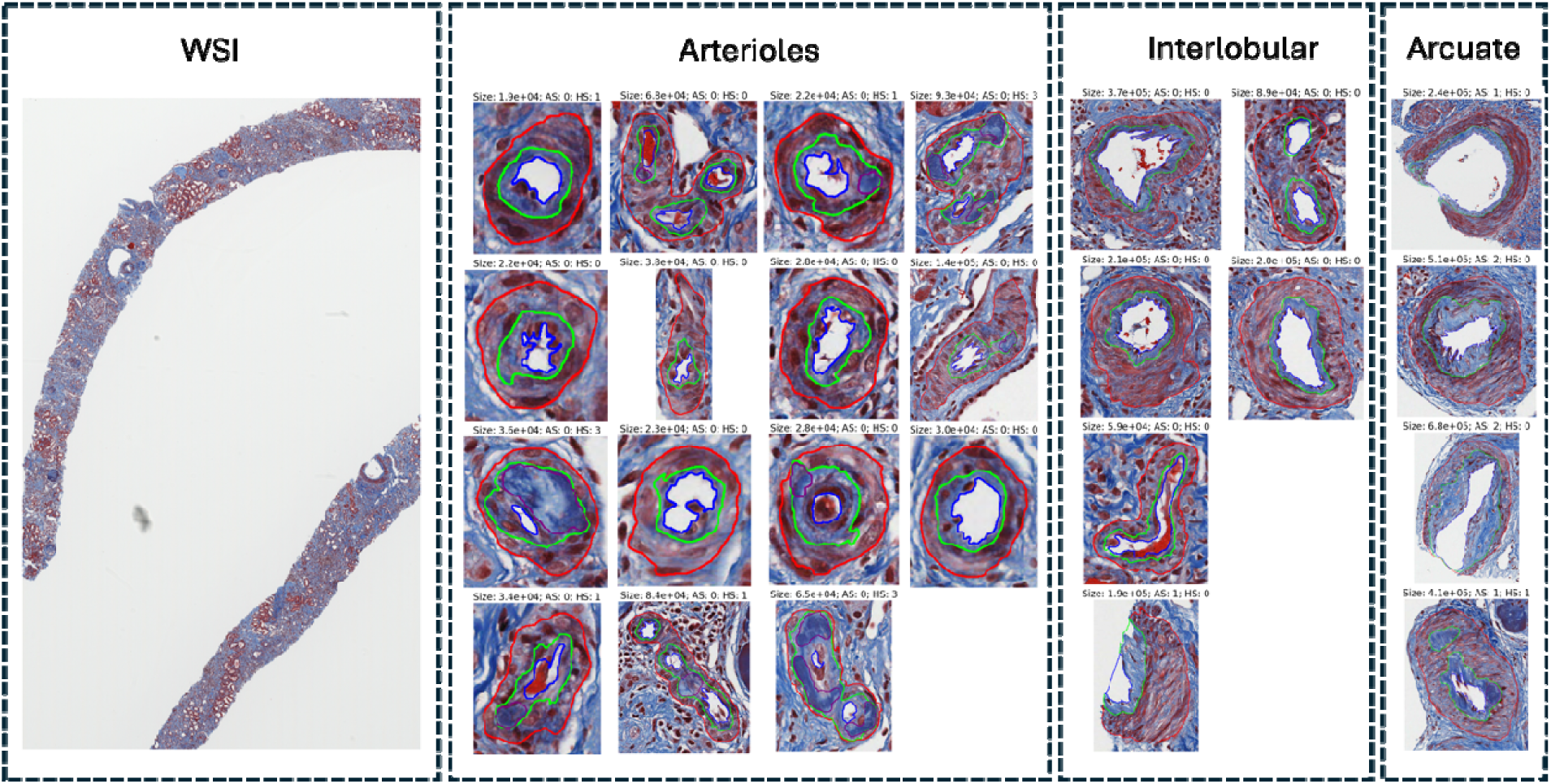
Arterial/Arteriolar Segmentation, Categorization and Scoring. Segmentation of muscular vessels and subcompartments, categorization of muscular vessels into arterioles and interlobular and arcuate arteries, and artery/arteriole-level visual scores for arteriosclerosis severity (AS) (0-3+) and arteriolar hyalinosis severity (HS) (0-3+) on a trichrome-stained whole slide image. The border of the arterial/arteriolar lumen is highlighted in blue, the outer border of the intima in green, the outer border of the media in red, and outer border of hyalinosis in purple. Displayed above each image is the size of each artery/arteriole in pixels, alongside the AS and HS scores.

Labelbox was also used for scoring each artery and arteriole for severities of the arteriosclerosis and hyalinosis using a semi-quantitative scale of 0 (none), 1 (mild, up to 25%), 2 (moderate, 25 to 50%), and 3 (severe, more than 50%).^36^ Scoring and categorization was performed by consensus of two study pathologists (Figure 2).

### Pathomic Quantification of Arterial/Arteriolar Characteristics

#### Area-based Feature Extraction

For each artery/arteriole, we calculated the area of the lumen, intima, media, and hyalinosis compartments, as well as the total artery/arteriole area. The area of each individual compartment was computed by counting the number of pixels within the segment of each compartment, while the total area of the arteries/arterioles is determined by summing the areas of the lumen, intima, and media compartments. To facilitate comparisons and analyses, we normalized the area measurements by representing the area of each compartment (lumen, intima, media, and hyalinosis) as a proportion of the total artery/arteriole area. Additionally, we transformed the artery/arteriole area using the logarithmic function to improve representation.

#### Thickness-based Feature Extraction

Using the segmentation boundaries of the lumen, intima and media, intra-arterial/arteriolar thickness was measured using radial sampling and ray casting techniques. Both intima and media thickness were measured and represented as functions of spatially encoded polar coordinates along the entire arterial/arteriolar perimeter. Signal-smoothing techniques were employed to mitigate the impact of variable arterial/arteriolar morphology caused during tissue harvesting and preparation (Figure 3).^22^ In general, this radial sampling technique provides two key advantages: (i) it allows for the selective exclusion of corrupted radial samples in arteries/arterioles with irregular appearances, such as edge artifacts and arterial bifurcations, and (ii) it facilitates the identification of radial samples with thickness values potentially influenced by hyalinosis. Thickness measurements were represented by three distinct series, each corresponding to a specific attribute: intima thickness, media thickness, and the intima-to-media thickness ratio. From these measurements, we extracted pathomic features, including global features (average and median) that describe the overall distribution of intra-arterial/arteriolar thickness, and local features (peak height and peak prominence) that highlight local variations in arterial/arteriolar thickness.

**Figure 3.**
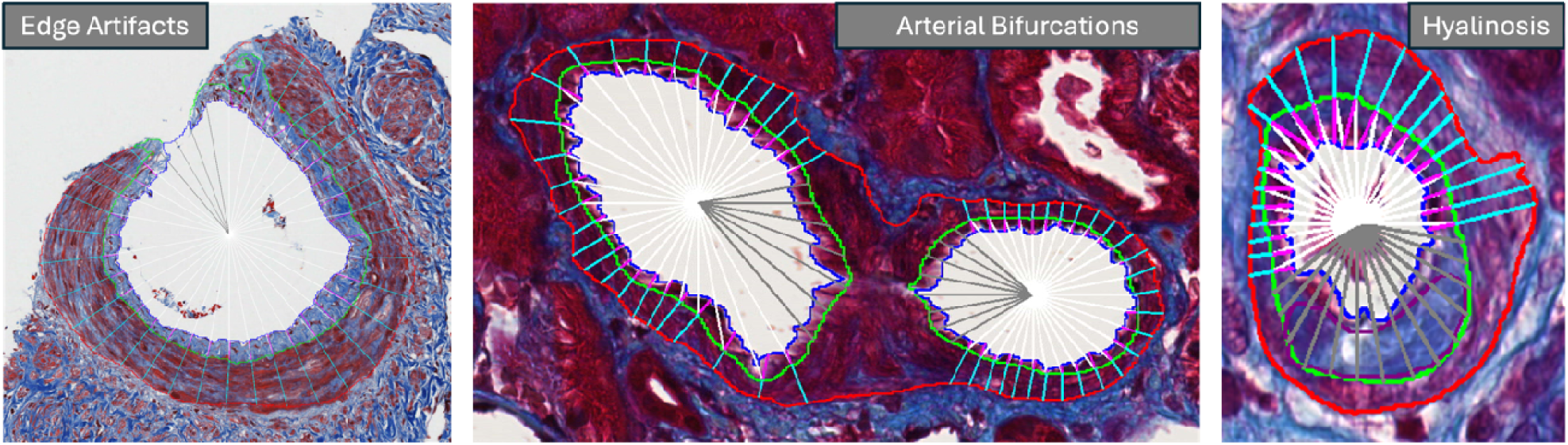
Intra-arterial/arteriolar thickness measurements based on radial sampling in arteries/arterioles with various appearances. Arcuate artery with edge artifacts (left column): Radial samples pointing to the open lumen are excluded for analysis (gray). For each of the remaining radial samples (white), the lengths of line segments are measured as thickness values for the intima (pink) and media (light blue). Interlobular artery with arterial bifurcations (middle column): Radial samples intersecting with other lumens or intima are discarded. Arterioles with hyalinosis (Right Panel): Radial samples intersecting with hyalinosis areas are discarded. Note: For all discarded samples in each case, adjacent samples within a predefined window are also discarded.

#### Statistical Analyses

We conducted the statistical analyses in two stages. First, at the artery/arteriole-level, we performed individual feature correlation analysis between pathomic and visual semiquantitative visual scoring. Second, we aggregated the artery/arteriole-level features and semiquantitative visual scores to the trichrome-stained WSI level and investigated their clinical relevance.

#### Association Between Pathomic Features and Semiquantitative Visual Scores at the Artery/Arteriole-Level

We investigated the association between the pathomic features extracted from each artery/arteriole and the corresponding artery/arteriole level semiquantitative visual scores for the arteriosclerosis and hyalinosis severities. Specifically, we examined the correlation between hyalinosis severity and hyalinosis area ratio, as well as the correlations between arteriosclerosis severity and various arterial and arteriolar features, including area measurements (area ratios of the lumen, intima, media, and logarithmically transformed total artery/arteriole area) and both global and local features extracted from thickness measurements of intima, media, and the intima-to-media ratio (Table 1). Spearman correlation coefficients were used to quantify the strength and direction of these relationships.

**Table 1.** Extracted artery/arteriole-level pathomic features for characterizing arteriosclerosis and hyalinosis.

| Feature Type | Feature |
| --- | --- |
| **Characterizing Hyalinosis** |  |
| Area-Based | Hyalinosis Area Ratio |
| **Characterizing Arteriosclerosis** |  |
| Area-Based | Log Artery Area |
|  | Media Area Ratio |
|  | Intima Area Ratio |
|  | Lumen Area Ratio |
| Thickness-Based | Intima Thickness Average |
|  | Intima Thickness Median |
|  | Intima Thickness Peak Height |
|  | Intima Thickness Peak Prominence |
|  | Media Thickness Average |
|  | Media Thickness Median |
|  | Media Thickness Peak Height |
|  | Media Thickness Peak Prominence |
|  | IM Thickness Ratio Average |
|  | IM Thickness Ratio Median |
|  | IM Thickness Ratio Peak Height |
|  | IM Thickness Ratio Peak Prominence |

Abbreviations: IM, intima-to-media.

Moreover, considering the interdependent relationships among the sixteen features used for characterizing arteriosclerosis (Table 1), we employed a decision tree model to evaluate feature importance, thereby identifying the most informative features that contribute to arteriosclerosis. This model measures the importance of a feature based on the decrease in node impurity, such as Gini impurity, achieved when the feature is used to split the data. The decision tree model was chosen for its ability to effectively handle non-linear relationships and interactions between multiple variables. This step is crucial for reducing dimensionality and selecting relevant features before aggregating data from the artery/arteriole level to the WSI level.

#### Prognostic Value of Arterial/Arteriolar Pathomic Features

To test the prognostic value of artery/arteriole-derived pathomic features and semiquantitative visual scores obtained for each artery/arteriole individually, we first aggregated them at the WSI-level by using descriptive statistics such as maximum, median, and 75th percentile. The clinical outcome of interest was a composite of disease progression outcomes, defined by the time from biopsy to a 40% decline in eGFR or kidney failure. To assess the prognostic value of these features, we employed multivariable Cox proportional hazards modeling. To mitigate the risk of overfitting, a least absolute shrinkage and selection operator (LASSO)-penalized Cox model was initially applied to select the most relevant features.

To individually test the prognostic value of different feature sets, we developed four distinct models, each incorporating unique inputs: 1) Demographics (age, sex, race, Hispanic ethnicity) and clinical characteristics (FSGS vs MCD, eGFR, and UPCR at the time of biopsy); 2)Biopsy-level semiquantitative visual scores: Retrieved from the NEPTUNE/CureGN core scoring dataset; 3)Trichrome-stained WSI-level semiquantitative visual scores: Derived by aggregating visual scores of individual arteries/arterioles within a single trichrome-stained WSI; 4) Trichrome-stained WSI-level pathomic features: Extracted at the individual artery/arteriole level, then aggregated to represent a single trichrome-stained WSI.

We considered the model with inputs consisting of only demographics and clinical characteristics as the baseline model. Building upon this baseline model, we developed three extended models, each integrating additional feature sets: 1) Baseline + Biopsy-level semiquantitative visual scores; 2) Baseline + Trichrome-stained WSI-level semiquantitative visual scores; and 3) Baseline + Trichrome-stained WSI-level pathomic features.

The concordance index (C-index), which measures the predictive power of the model, was used to evaluate the performance across three different artery/arteriole types. Additionally, we evaluated the association between each selected trichrome-stained WSI-level pathomic feature and semiquantitative visual score, identified through the LASSO-penalized Cox model, and the clinical outcome using separate standard Cox proportional hazards regression models. These evaluations were reported under two scenarios: unadjusted and adjusted for demographics and clinical characteristics.

## Results

### Study Cohorts and Sample Selection

Using the inclusion criteria described in the methods section, 127 FSGS and 98 MCD, for a total of 225 cases (1 trichrome-stained WSI per case), were included for analysis. The demographics and clinical data at the time of biopsy are summarized in Table 2.

**Table 2:** Summary of demographics and clinical data at the time of biopsy. Values are presented as count (percentage) for categorical variables and median (interquartile range) for continuous variables.

| **Variable** | **Study Sample (N=225)** |
| --- | --- |
| **Diagnosis** |  |
| MCD | 98 (43.56%) |
| FSGS | 127 (56.44%) |
| **Demographics** |  |
| Male sex | 130 (57.78%) |
| Age at biopsy, younger than 18 yr | 106 (47.1%) |
| **Race** |  |
| Asian or Asian American | 23 (10.22%) |
| Black or African American | 59 (26.22%) |
| White or Caucasian | 118 (52.44%) |
| Others (multiracial or unknown) | 25 (11.11%) |
| **Hispanic Ethnicity** |  |
| Hispanic or Latino | 53 (23.56%) |
| **Baseline Laboratory Values** |  |
| eGFR (ml/min per 1.73 m<sup>2</sup>) | 84.76 (54.11–105.69) |
| UPCR (g/g) | 3.20 (1.15–8.67) |
| **Arteriosclerosis Descriptor Score** |  |
| Absent | 120 (53.33%) |
| Mild (1-25%) | 46 (20.44%) |
| Moderate (26-50%) | 19 (8.44%) |
| Severe (>50%) | 13 (5.78%) |
| Unknown | 27 (12.00%) |
| **Hyalinosis Descriptor Score** |  |
| Absent | 150 (66.67%) |
| Mild (1-25%) | 40 (17.78%) |
| Moderate (26-50%) | 11 (4.89%) |
| Unknown | 24 (10.67%) |

Abbreviations: eGFR, estimated glomerular filtration rate; UPCR, urine protein-to-creatinine ratio; MCD, minimal change disease; FSGS, focal segmental glomerulosclerosis.

### Arterial/Arteriolar Segmentation, Categorization and Scoring

The distribution of arteries and arterioles varied across WSIs. In total, we segmented 1,499 arterioles across 217/225 WSIs (96.4% of total WSIs), 686 interlobular arteries across 196/225 WSIs (87.1% of total WSIs), and 131 arcuate arteries across 84/225 WSIs (37.3% of total WSIs). An additional 583 muscular vessels across 156/225 WSIs (69.3% of total WSIs) were excluded due to having only a single muscular layer, an absence of lumen, or the presence of artifacts.

Arterioles exhibited a high frequency of hyalinosis, with 284 (18.9%) arterioles showing hyalinosis compared to 31 (5.0%) interlobular and 1 (0.8%) arcuate artery. In contrast, arteriosclerosis was less frequent in arterioles (74 – 4.9%), but more prevalent in interlobular arteries (152 – 22.2%) and arcuate arteries (79 – 60.3%). (Table 3)

**Table 3.** Distribution of artery/arteriole level visual scores of arteriosclerosis and hyalinosis.

| **Artery/Arteriole Type** | **Arterioles**<br>**(n=217)** | **Interlobular Arteries**<br>**(n=196)** | **Arcuate Arteries**<br>**(n=84)** |
| --- | --- | --- | --- |
| Total Count | 1,499 | 686 | 131 |
| **Arteriosclerosis Severity Score** |  |  |  |
| Score 0 | 1,425 | 534 | 52 |
| Score 1 | 60 | 92 | 34 |
| Score 2 | 11 | 31 | 27 |
| Score 3 | 3 | 29 | 18 |
| **Hyalinosis Severity Score** |  |  |  |
| Score 0 | 1,215 | 652 | 130 |
| Score 1 | 220 | 31 | 1 |
| Score 2 | 47 | 3 | 0 |
| Score 3 | 17 | 0 | 0 |

The number of WSIs containing each vessel type is denoted by (n).

### Artery/Arteriole-Level Pathomic Feature Analysis

#### Hyalinosis

The hyalinosis area ratio demonstrated a moderate correlation with severity scores in arterioles (p=0.46, p<0.001). In contrast, the correlation was low in interlobular arteries (p=0.04, p=0.30), suggesting limited predictive value of this feature. Due to the rarity of hyalinosis in arcuate arteries, with only one instance observed, statistical correlation analysis was not informative. (Supplementary Figure 1)

#### Arteriosclerosis

In arterioles, both area and thickness measurements demonstrate a weak correlation with arteriosclerosis severity (p=0.27, p<0.001) (Supplementary Figure 2). In contrast, in interlobular and arcuate arteries, multiple area and thickness pathomic features were significantly correlated with arteriosclerosis semiquantitative visual scores. Notably, the feature of the intima-media ratio average exhibited a strong correlation in interlobular arteries (p=0.69, p<0.001) and in arcuate arteries (p=0.80, p<0.001) (Supplementary Figure 3 and 4.). In our decision tree model aimed at identifying the most informative features contributing to arteriosclerosis, we observed distinct patterns across different artery/arteriole types. (Table 4) For both interlobular and arcuate arteries, features such as the “intima area ratio” and the “intima-media (IM) thickness ratio” (average or median) contributed significantly. In arterioles, in addition to the “intima area ratio” and “intima thickness median,” the “log artery/arteriole area” and “lumen area ratio” features also demonstrated high importance.

**Table 4.** Selected features for characterizing arteriosclerosis for different artery/arteriole types derived from a decision tree model.

| Artery/Arteriole Type | Feature Name | Importance |
| --- | --- | --- |
| Arterioles | Intima Area Ratio | 0.246 |
|  | Log Artery / Arteriole Area | 0.185 |
|  | Lumen Area Ratio | 0.129 |
|  | Intima Thickness Median | 0.104 |
| Interlobular Arteries | IM Thickness Ratio Average | 0.461 |
|  | Intima Area Ratio | 0.212 |
| Arcuate Arteries | Intima Area Ratio | 0.309 |
|  | IM Thickness Ratio Median | 0.207 |

Abbreviations: IM, intima-to-media.

Therefore, the “hyalinosis area ratio” feature characterizing hyalinosis and selected features characterizing arteriosclerosis (Table 4) are aggregated at the WSI-level for clinical outcome analysis.

#### Clinical Outcome of Patient-level Pathomic Features Analysis and Semiquantitative Visual Scoring

Among all individual parameter sets, the model using demographics and clinical characteristics alone achieved the highest performance to predict the composite outcome of 40% decline in eGFR or kidney failure across all three artery/arteriole types, with C-indices of 0.70 for arterioles and interlobular arteries, and 0.69 for arcuate arteries. Both WSI-level visual scores and WSI-level pathomic features consistently outperformed biopsy-level visual scores. Notably, for arterioles, WSI-level pathomic features (C-index: 0.68) demonstrated superior performance compared to WSI-level visual scores (C-index: 0.66). (Table 5)

**Table 5.** Concordance index (C-index) for cox models by artery/arteriole type, for individual and combined parameter sets.

| Model Parameters | Arterioles<br>(n=192) | Interlobular<br>(n=174) | Arcuate<br>(n=71) |
| --- | --- | --- | --- |
| Demographics and Clinical Characteristics<br>(Baseline) | 0.70 | 0.70 | 0.69 |
| **Individual Parameter Set** |  |  |  |
| Biopsy-Level Visual Scores | 0.59 | 0.58 | 0.56 |
| WSI-Level Visual Scores | 0.66 | 0.61 | 0.67 |
| WSI-Level Pathomic Features | 0.68 | 0.60 | 0.59 |
| **Combined Parameter Set** |  |  |  |
| Baseline + Biopsy-Level Visual Scores | 0.70 | 0.72 | 0.72 |
| Baseline + WSI-Level Visual Scores | 0.73 | 0.71 | 0.75 |
| Baseline + WSI-Level Pathomic Features | 0.75 | 0.72 | 0.74 |

The patient count refers to patients with complete data across all categories: demographics and clinical characteristics, biopsy-level visual scores, trichrome-stained WSI-level visual scores, and trichrome-stained WSI-level pathomic features. The number of patients per artery/arteriole type is denoted by (n).

**Table 6.** Associations between individual trichrome-stained WSI-level semiquantitative visual scores and clinical outcomes from Cox proportional hazards models.

| Feature | Model | HR (95% CI) | p-value |
| --- | --- | --- | --- |
| *Arterioles* |  |  |  |
| 75th Hyalinosis Severity | Unadjusted | 1.06 (0.80-1.40) | 0.693 |
|  | Adjusted | 1.01 (0.74-1.38) | 0.949 |
| **Median Hyalinosis Severity** | **Unadjusted** | **1.26 (1.01-1.57)** | **0.043** |
|  | Adjusted | 1.26 (0.99-1.60) | 0.057 |
| Max Hyalinosis Severity | Unadjusted | 1.24 (0.92-1.66) | 0.154 |
|  | Adjusted | 1.23 (0.87-1.73) | 0.238 |
| **Max Arteriosclerosis Severity** | **Unadjusted** | **1.32 (1.05-1.65)** | **0.016** |
|  | **Adjusted** | **1.32 (1.04-1.67)** | **0.023** |
| *Interlobular Arteries* |  |  |  |
| Median Arteriosclerosis Severity | Unadjusted | 0.97 (0.72-1.32) | 0.861 |
|  | Adjusted | 0.88 (0.63-1.25) | 0.487 |
| Max Arteriosclerosis Severity | Unadjusted | 1.07 (0.81-1.42) | 0.629 |
|  | Adjusted | 1.02 (0.73-1.41) | 0.928 |
| *Arcuate Arteries* |  |  |  |
| Median Arteriosclerosis Severity | Unadjusted | 0.82 (0.52-1.31) | 0.409 |
|  | Adjusted | 0.68 (0.34-1.39) | 0.291 |
| Max Arteriosclerosis Severity | Unadjusted | 0.95 (0.59-1.53) | 0.828 |
|  | Adjusted | 0.87 (0.48-1.57) | 0.645 |

Arteriosclerosis and hyalinosis severities were initially visually scored at the artery/arteriole level, aggregated to represent a single trichrome-stained WSI (using median, 75th percentile, and maximum values—denoted as prefixes for each feature name).

**Table 7.** Associations between individual trichrome-stained WSI-level pathomic features and clinical outcomes from Cox proportional hazards models.

| Feature | Model | HR (95% CI) | p-value |
| --- | --- | --- | --- |
| *Arterioles* |  |  |  |
| 75th Log Artery / Arteriole Area | Unadjusted | 1.00 (0.99-1.01) | 0.999 |
|  | Adjusted | 1.00 (1.00-1.00) | 0.999 |
| **75th Lumen Area Ratio** | Unadjusted | 0.73 (0.53-1.01) | 0.057 |
|  | **Adjusted** | **0.69 (0.48-0.98)** | **0.037** |
| 75th Intima Thickness Median | Unadjusted | 1.31 (0.99-1.74) | 0.063 |
|  | Adjusted | 1.30 (0.96-1.78) | 0.093 |
| Median Intima Thickness Median | Unadjusted | 1.11 (0.81-1.53) | 0.519 |
|  | Adjusted | 1.09 (0.77-1.55) | 0.631 |
| Max Log Artery / Arteriole Area | Unadjusted | 1.14 (0.83-1.55) | 0.416 |
|  | Adjusted | 1.15 (0.82-1.60) | 0.419 |
| Median Log Artery / Arteriole Area | Unadjusted | 1.04 (0.77-1.40) | 0.804 |
|  | Adjusted | 1.02 (0.74-1.41) | 0.908 |
| **Max Hyalinosis Area Ratio** | Unadjusted | 1.24 (0.99-1.55) | 0.061 |
|  | **Adjusted** | **1.28 (1.00-1.62)** | **0.046** |
| *Interlobular Arteries* |  |  |  |
| 75th IM Thickness Ratio Average | Unadjusted | 0.95 (0.70-1.30) | 0.742 |
|  | Adjusted | 0.92 (0.65-1.29) | 0.617 |
| Max Intima Area Ratio | Unadjusted | 1.08 (0.81-1.44) | 0.579 |
|  | Adjusted | 1.04 (0.73-1.46) | 0.841 |
| Median Intima Area Ratio | Unadjusted | 1.00 (0.74-1.37) | 0.975 |
|  | Adjusted | 1.00 (1.00-1.00) | 0.999 |
| Median IM Thickness Ratio Average | Unadjusted | 0.95 (0.69-1.29) | 0.725 |
|  | Adjusted | 0.91 (0.65-1.28) | 0.589 |
| *Arcuate Arteries* |  |  |  |
| 75th Intima Area Ratio | Unadjusted | 0.69 (0.41-1.16) | 0.163 |
|  | Adjusted | 0.54 (0.27-1.09) | 0.084 |
| Median IM Thickness Ratio Median | Unadjusted | 0.71 (0.43-1.17) | 0.174 |
|  | Adjusted | 0.57 (0.29-1.12) | 0.103 |

Pathomic features were extracted at the individual artery/arteriole level and then aggregated to represent a single trichrome-stained WSI (using median, 75th percentile, and maximum values—denoted as prefixes for each feature name).

Compared to using demographics and clinical characteristics only, the addition of biopsy-level semiquantitative visual scores from the NEPTUNE/CureGN core scoring data marginally increased the C-index in arcuate arteries from 0.69 to 0.72 and in interlobular arteries from 0.70 to 0.72, indicating a limited improvement in performance. Moreover, this enhancement was not observed in arterioles, where the C-index remained unchanged at 0.70. The addition of either trichrome-stained WSI-level semiquantitative visual scores or pathomic features to arterioles increased the C-index from 0.70 with only demographics and clinical characteristics to 0.73 with trichrome-stained WSI-level semiquantitative visual scores and 0.75 with trichrome-stained WSI-level pathomic features. Similarly, in arcuate arteries, the C-index improved from 0.69 to 0.75 with trichrome-stained WSI-level semiquantitative visual scores and 0.74 with trichrome-stained WSI-level pathomic features. In contrast, for interlobular arteries, the C-index changes were minimal, moving from 0.70 with only demographics and clinical characteristics to 0.71 with trichrome-stained WSI-level semiquantitative visual scores and to 0.72 with the addition of trichrome-stained WSI-level pathomic features. (Table 5)

Our analysis revealed no significant associations between trichrome-stained WSI-level semiquantitative visual scores or pathomic features and clinical outcomes in arcuate and interlobular arteries (Tables 6 & 7 respectively). However, in arterioles, several trichrome-stained WSI-level semiquantitative visual scores and pathomic features demonstrated significant associations with clinical outcomes, both before and after adjustment. Notably, for trichrome-stained WSI-level semiquantitative visual scores, both unadjusted and adjusted ‘max arteriosclerosis severity’ exhibited a significant HR of 1.32 (p < 0.05), while the unadjusted ‘median hyalinosis severity’ demonstrated an HR of 1.26 (p < 0.05). Among trichrome-stained WSI-level pathomic features, the adjusted ‘75th percentile lumen area ratio’ in arterioles showed a hazard ratio (HR) of 0.69 (p < 0.05), while the adjusted ‘max hyalinosis area ratio’ showed an HR of 1.28 (p < 0.05), indicating their relevance in predicting clinical outcomes

## Discussion

In this study, we developed a computational approach to segment muscular vessels and their sub-compartments (media, intima, lumen) in digital kidney biopsies of patients with FSGS and MCD, and to computationally characterize viable arteries/arterioles using pathomic features extracted from the segmented sub-compartments. We then demonstrated the prognostic value of the extracted pathomic features to predict the composite outcome of 40% decline in eGFR or kidney failure compared to conventional methodologies.

When the analysis was conducted at the patient level, both trichrome-stained WSI-level pathomic features and semiquantitative visual scores demonstrated better performance than biopsy-level visual scores, indicating the value of WSI-derived data in capturing artery/arteriole-level information. WSI-level pathomic features (C-index: 0.68) exhibited better performance than WSI-level visual scores in arterioles, underscoring the potential of pathomic features in enhancing prognostic models.

When added to baseline models containing demographics and clinical characteristics, both trichrome-stained WSI-level pathomic features and semiquantitative visual scores, particularly in arterioles and arcuate arteries, enhanced predictive performance. Notably, trichrome-stained WSI-level pathomic features and semiquantitative visual scores proved more effective in predicting clinical outcomes than biopsy-level visual scores (assessed across all stains of the biopsy). This increased effectiveness may be attributed to an increased robustness and consistency of data obtained by aggregating at trichrome-stained WSI-level computer-based pathomic features extracted from each artery/arteriole and the semiquantitative visual scores obtained by consensus from artery/arteriole, respectively.

Additionally, pathomic features extracted from arterioles, such as the ‘75th percentile lumen area ratio’ and the ‘max hyalinosis area ratio ‘— along with corresponding visual scores like ‘max arteriosclerosis severity’ and ‘median hyalinosis severity’ — showed independent associations with clinical outcomes. The selected trichrome-stained WSI-level pathomic features and semiquantitative visual scores are highly relevant, as hyalinosis commonly occurs in arterioles, with severity being correlated to its area ratio, and the lumen area ratio serves as a key indicator of arteriosclerosis. Moreover, the use of pathomic features offers additional advantages in terms of reproducibility and efficiency, as they are less prone to variability in interpretation and can be automated, making them a reliable tool in clinical settings. No individual pathomic features or semiquantitative scores were selected for interlobular and arcuate arteries, likely due to the larger number arterioles present in our dataset compared to interlobular and arcuate arteries.

Compared to previous work on arterial features demonstrating the prognostic value of scoring arteriosclerosis and arteriolar hyalinosis ^22,2537,38^, this study offers several advancements. First, we developed a highly detailed dataset, comprised of three sets of severity scores for arteriosclerosis and hyalinosis: (1) biopsy-level semiquantitative visual scores using all WSIs available, (2) individual artery/arteriole semiquantitative visual scores, and (3) aggregated semiquantitative visual scores at the trichrome WSI level. Second, our application of computational approaches enables a precise measurement of parameters and the extraction of quantifiable data, which may enhance diagnostics compared to visual representation. The application of pathomic features such as area-based features and intra-arterial thickness features, therefore have the potential of increasing reproducibility, efficiency, and clinical relevance of measurable vascular attributes.

This study has several limitations. First, the dataset size is limited, especially when evaluating the prognostic values of pathomic features and visual scores for arcuate arteries, where only 70 patients were utilized. Future studies will seek to expand the dataset by incorporating more FSGS/MCD cases as well as other diseases, such as diabetic nephropathy, IgA nephropathy, and hypertension, which could enhance the generalizability of the findings. Second, the segmentation of muscular vessels and their sub-compartments (media, intima, lumen) requires manual quality control prior to pathomic feature extraction. This is largely due to the variability in the image presentation such as cross section of the artery, variability in size and muscular or intima thickness within each vessel type and within the same vessel. However, by incorporating additional datasets, we anticipate improving the generalizability of the segmentation models.

In conclusion, we developed a computational framework for robust and reproducible characterization of arterial/arteriolar injury that can enhance our ability to prognosticate and predict disease progression in patients with MCD and FSGS and that can ultimately be applied, with careful testing for generalizability and validation, in clinical research and practice.

## Data Availability

All data produced in the present work are contained in the manuscript

## Acknowledgments

NEPTUNE Collaborating Sites: *Atrium Health Levine Children’s Hospital, Charlotte, SC*: Susan Massengill*, Layla Lo^#^ *Cleveland Clinic, Cleveland, OH*: Katherine Dell*, John O’Toole*, John Sedor**, Victoria Grange^#^ *Children’s Hospital, Denver, CO:* Bradley Dixon*, Nathan Rogers^#^

*Children’s Hospital, Los Angeles, CA*: Rachel Lestz*, Natalie Esquivias^#^

*Children’s Mercy Hospital, Kansas City, MO*: Tarak Srivastava*, Kelsey Markus^#^ *Cohen Children’s Hospital, New Hyde Park, NY*: Christine Sethna*, Suzanne Vento^#^ *Columbia University, New York, NY:* Pietro Canetta*

*Duke University Medical Center, Durham, NC:* Opeyemi Olabisi*, Rasheed Gbadegesin**, Maurice Smith^#^

*Emory University, Atlanta, GA:* Laurence Greenbaum*, Chia-shi Wang*, Chris Fan^#^

*The Lundquist Institute, Torrance, CA:* Sharon Adler*, Janine LaPage^#^

*John H Stroger Cook County Hospital, Chicago, IL:* Amatur Amarah*

*Johns Hopkins Medicine, Baltimore, MD:* Meredith Atkinson*, Ryan Hutson^#^

*Mayo Clinic, Rochester, MN:* John Lieske, Marie Hogan, Fernando Fervenza

*Medical University of South Carolina, Charleston, SC:* David Selewski*, Cheryl Alston^#^

*Montefiore Medical Center, Bronx, NY:* Kim Reidy*, Michael Ross*, Frederick Kaskel**, Patricia Flynn^#^

*New York University Medical Center, New York, NY:* Laura Malaga-Dieguez*, Olga Zhdanova**, Laura Jane Pehrson^#^, Melanie Miranda^#^

*The Ohio State University College of Medicine, Columbus, OH*: Salem Almaani*, Laci Roberts^#^ *Riley Children’s Hospital of Indiana University, Indianapolis, IN:* Myda Khalid*, Veronica Servin^#^ *Stanford University, Stanford, CA:* Richard Lafayette*, Elizabeth Chen^#^

*Temple University, Philadelphia, PA:* Iris Lee**

*Texas Children’s Hospital at Baylor College of Medicine, Houston, TX*: Shweta Shah*, Thinh Phan^#^

*University Health Network Toronto:* Heather Reich*, Michelle Hladunewich**, Paul Ling^#^, Martin Romano^#^

*University of California at San Diego, San Diego, CA:* Ambarish Athavale*, Caitlin Carter*, Kristin Zeeb^#^

*University of California at San Francisco, San Francisco, CA*: Paul Brakeman*, Daniel Schrader

*University of Colorado Anschutz Medical Campus, Aurora, CO*: James Dylewski* Nathan Rogers^#^

*University of Kansas Medical Center, Kansas City, KS*: Ellen McCarthy*, Catherine Creed^#^

*University of Miami, Miami, FL:* Alessia Fornoni*, Miguel Bandes^#^

*University of Michigan, Ann Arbor, MI:* Matthias Kretzler*, Laura Mariani*, Zubin Modi*, Amanda Williams^#^, Roxy Ni^#^

*University of Minnesota, Minneapolis, MN:* Patrick Nachman*, Michelle Rheault*, Ariel Langenberger^#^, Brady Wallner^#^

*University of North Carolina, Chapel Hill, NC:* Vimal Derebail*, Keisha Gibson*, Anne Froment^#^, Sharia Warren^#^

*University of Pennsylvania, Philadelphia, PA:* Lawrence Holzman*, Kevin Meyers**, Krishna Kallem^#^, Arielle Swenson^#^

*University of Texas San Antonio, San Antonio, TX*: Samin Sharma**

*University of Texas Southwestern, Dallas, TX:* Elizabeth Roehm*, Kamalanathan Sambandam**, Elizabeth Brown**

*University of Washington, Seattle, WA:* Ashley Jefferson*, Sangeeta Hingorani**, Katherine Tuttle^**§^, Linda Manahan ^#^, Emily Pao^#^, Kelli Kuykendall^§^

*Wake Forest University Baptist Health, Winston-Salem, NC:* Jen Jar Lin**

*Washington University in St. Louis, St. Louis, MO*: Brian Stotter*, Joseph Dumayas^#^

## Data Analysis and Coordinating Center

*University of Michigan:* Matthias Kretzler*, Brenda Gillespie**, Laura Mariani**, Zubin Modi**, Eloise Salmon**, Howard Trachtman**, Tina Mainieri, Michael Arbit, Hailey Desmond, Sean Eddy, Damian Fermin, Wenjun Ju, Maria Larkina, Chrysta Lienczewski, Rebecca Scherr, Jonathan Troost, Amanda Williams, Yan Zhai;; *Cleveland Clinic:* Crystal Gadegbeku**, John Sedor**, *Duke University:* Laura Barisoni**; *Harvard University:* Matthew G Sampson**; *Northwestern University:* Abigail Smith**; *University of Pennsylvania:* Lawrence Holzman**, Jarcy Zee**

## Digital Pathology Committee

Carmen Avila-Casado *(University Health Network)*, Serena Bagnasco *(Johns Hopkins University)*, Lihong Bu *(Mayo Clinic)*, Shelley Caltharp *(Emory University)*, Clarissa Cassol *(Arkana)*, Dawit Demeke *(University of Michigan)*, Brenda Gillespie *(University of Michigan)*, Jared Hassler *(Temple University)*, Leal Herlitz *(Cleveland Clinic)*, Stephen Hewitt *(National Cancer Institute)*, Jeff Hodgin *(University of Michigan)*, Danni Holanda *(Arkana)*, Neeraja Kambham *(Stanford University)*, Kevin Lemley, Laura Mariani *(University of Michigan)*, Nidia Messias *(Washington University)*, Alexei Mikhailov *(Wake Forest)*, Vanessa Moreno *(University of North Carolina)*, Behzad Najafian *(University of Washington)*, Matthew Palmer *(University of Pennsylvania)*, Avi Rosenberg *(Johns Hopkins University)*, Virginie Royal *(University of Montreal)*, Miroslav Sekulik *(Columbia University)*, Barry Stokes *(Columbia University)*, David Thomas *(Duke University)*, Ming Wu *(University of New York)*, Michifumi Yamashita *(Cedar Sinai)*, Hong Yin *(Emory University)*, Jarcy Zee *(University of Pennsylvania)*, Yiqin Zuo *(University of Miami)*. Co-Chairs: Laura Barisoni *(Duke University)*, Cynthia Nast *(Cedar Sinai)*.

## CureGN Collaborators

The CureGN Consortium members listed below, from within the four Participating Clinical Center networks and Data Coordinating Center, are acknowledged by the authors as Collaborators.

**CureGN Principal Investigators; *CureGN Site Principal Investigators; ^+^CureGN Pathologists,

^#^CureGN Lead Coordinators.

### CureGN Participating Clinical Centers (PCC) through Columbia University

*Columbia University, New York, NY, US*: Gerald Appel, Revekka Babayev, Ibrahim Batal ^+^, Andrew Bomback**, Pietro Canetta, Brenda Chan, Vivette Denise D’Agati ^+^, Samitri Dogra, Hilda Fernandez, Gabriele Gaggero^+^, Ali Gharavi**, William Hines, Syed Ali Husain, Krzysztof Kiryluk**, Satoru Kudose ^+^, Fangming Lin, Victoria Kolupaeva#, Maddalena Marasa, Glen Markowitz ^+^, Mariela Naarro-Torres, Hila Milo Rasouly, Sumit Mohan, Nicola Mongera, Jordan Nestor, Thomas Nickolas, Jai Radhakrishnan, Maya Rao, Maya Sabatello, Simone Sanna-Cherchi, Dominick Santoriello^+^, Miroslav Sekulic ^+^, Shayan Shirazian, Michael Barry Stokes^+^, Natalie Uy, Natalie Vena, Benjamin Wooden

*University of Warsaw, Warszawa, Poland:* Bartosz Foroncewicz, Natalia Krata, Barbara Moszczuk, Krzysztof Mucha*, Agnieszka Perkowska-Ptasińska, Elżbieta Ryszkowska

*IRCCS Giannina Gaslini, Genoa, Italy:* Gian Marco Ghiggeri*, Francesca Lugani, Valerio Vellone^+^

### CureGN Participating Clinical Centers (PCC) through the Pediatric Nephrology Research Consortium

*Children’s Hospital of Michigan, Detroit, MI, USA*: Rossana Baracco, Amrish Jain* *Children’s Hospital of New Orleans/ LSU Health, New Orleans, LA, USA*: Diego Aviles* *Children’s Mercy Hospital, Kansas City, MO, USA*: Tarak Srivastava*, Alexander Katz^+^ *Children’s National Medical Center, Wmcgashington DC, USA*: Sun-Young Ahn*

*Cincinnati Children’s Hospital Cincinnati, OH, USA*: Prasad Devarajan, Elif Erkan*, Donna Claes, Hillarey Stone

*Connecticut Children’s Medical Center, Hartford, CT, USA*: Sherene Mason*

*East Carolina University Brody School of Medicine, Greenville, NC, USA*: Liliana Gomez-Mendez*

*Emory University, Atlanta, GA, USA*: Larry Greenbaum**, Chia-shi Wang, Hong (Julie) Yin^+^ *Helen DeVos Children’s Hospital, Grand Rapids, MI, USA*: Goebel Jens*, Julia Steinke *Levine Children’s Hospital/Atrium Health, Charlotte, NC, USA*: Donald Weaver*

*Lurie Children’s Hospital, Chicago IL, USA*: Jerome Lane*

*Mayo Clinic, Rochester, MN, USA*: Carl Cramer*

*Medical College of Wisconsin, Milwaukee, WI, USA*: Cindy Pan, Neil Paloian, Rajasree Sreedharan*

*Medical University of South Carolina, Charleston SC, USA*: David Selewski, Katherine Twombley*, Sally Self^+^

*Nationwide Children’s Hospital, Columbus, OH, USA*: Samantha Martinek-Bundt#, Dawson Carmean#, Mary Dreher^#^, Aria Dockham^#^, Mahmoud Kallash*, John Mahan, Samantha Sharpe^#^, William Smoyer**, Laura Biederman^+^

*Oregon Health and Science University, Portland, OR, USA*: Amira Al-Uzri*, Sandra Iragorri

*Riley Children’s Hospital, Indianapolis, IN, USA*: Myda Khalid**

*Cardinal Glennon Children’s Medical Center/ St. Louis University, St. Louis, MO, USA*: Craig Belsha*

*Texas Children’s Hospital, Houston, TX, USA*: Elizabeth Onugha*, Michael Braun, AC Gomez

*Texas Tech Health Sciences Center, Amarillo, TX, USA*: Tetyana Vasylyeva*

*Children’s of Alabama, University of Alabama, Birmingham, AL, USA*: Daniel Feig* *University of Colorado Children’s Hospital, Colorado, Aurora, CO, USA*: Melisha Hannah* *University of Iowa Children’s Hospital, Iowa City, IA, USA*: Carla Nester*

*University of Kentucky, Lexington, KY, USA*: Aftab Chishti*

*University of Louisville, Louisville, KY, USA*: Jon Klein**

*Holtz Medical Center, University of Miami, Miami, FL, USA*: Chryso Katsoufis, Wacharee Seeherunvong*

*University of Minnesota Children’s Hospital, Minneapolis, MN, USA*: Michelle Rheault** *University of New Mexico Health Sciences Center, Albuquerque, NM, USA*: Craig Wong* *University of Oklahoma Health Sciences Center, Oklahoma City, OK, USA*: Qassim Abid* *University of Virginia, Charlottesville, VA, USA*: John Barcia*, Agnes Swiatecka-Urban *University of Wisconsin, Madison, WI, USA*: Sharon Bartosh*

*Vanderbilt Children’s Hospital, Nashville TN, USA*: Tracy Hunley*

*Washington University in St. Louis, St. Louis, MO, USA*: Vikas Dharnidharka*, Brian Stotter, Joseph, Gaut ^+^

### CureGN Participating Clinical Centers (PCC) through the University of North Carolina

*Hôpital Maisonneuve-Rosemont, Montreal, Canada*: Louis-Philippe Laurin*, Virginie Royal^+^, Mathieu Latour^+^, Natlie (Natacha) Patey ^+^

*Medical University of South Carolina, Charleston, SC, USA*: Anand Achanti, Milos Budisavljevic*

*Northwestern University, Chicago, IL, USA*: Cybele Ghossein, Yonatan Peleg *

*Ohio State University, Columbus, OH, USA*: Salem Almaani*, Isabelle Ayoub, Samir Parikh, Brad Rovin, Anjali Satoskar^+^

*University of Chicago, Chicago, IL, USA*: Anthony Chang^+^

*University of Alabama at Birmingham, Birmingham, AL, USA*: Huma Fatima^+^, Jan Novak, Matthew Renfrow, Dana Rizk*

*<u>University of North Carolina Kidney Center, Chapel Hill, NC, USA</u>*: Dhruti Chen, Vimal Derebail**, Ronald Falk**, Keisha Gibson, Dorey Glenn, Susan Hogan, Koyal Jain, J. Charles Jennette^+^, Amy Mottl, Caroline Poulton^#^, Monica Reynolds, Manish Kanti Saha, Nicole E. Wyatt

*Vanderbilt University, Nashville, TN, USA*: Agnes Fogo^+^, Neil Sanghani*

*Virginia Commonwealth University, Richmond, VA, USA*: Jason Kidd*, Selvaraj Muthusamy^+^

### CureGN Participating Clinical Centers (PCC) through the University of Pennsylvania

*Children’s Hospital of Philadelphia, Philadelphia, PA, USA*: Michelle Denburg, Amy Kogon, Kevin Meyers*, Madhura Pradhan

*Cleveland Clinic, Cleveland, OH, CA*: Raed Bou Matar*, John O’Toole, John Sedor

Cohen Children’s Medical Center, New Hyde Park, NY, USA: Christine Sethna*^, Suzanne Vento^#^

*Johns Hopkins University, Baltimore, MD, USA*: Mohamed Atta, Serena Bagnasco^+^, Alicia Neu, John Sperati*

*Lundquist Institute at Harbor-UCLA Medical Center, Torrance, CA, USA*: Sharon Adler*, Tiane Dai, Ram Dukkipati

*Mayo Clinic, Rochester, MN, USA*: Fernando Fervenza*, Sanjeev Sethi ^+^

*Montefiore Medical Center, The Bronx, New York, NY, USA*: Frederick Kaskel, Kaye Brathwaite, Kimberly Reidy*

*New York University, New York, NY, USA*: Joseph Weisstuch, Ming Wu ^+^, Olga Zhdanova

*Spokane Providence Medical Center, Spokane, WA, USA*: Katherine Tuttle*

*Stanford University, Palo Alto, CA, USA*: Jill Krissberg, Richard Lafayette*, Kamal Fahmeedah, Elizabeth Talley

*Sunnybrook Health Sciences Centre, Toronto, Canada*: Michelle Hladunewich*

*The Hospital for Sick Children, Toronto, Canada*: Rulan Parekh*

*University Health Network, Toronto, Canada*: Carmen Avila-Casado^+^, Daniel Cattran*, Reich Heather, Philip Boll

*University of Miami, Miami, FL, USA*: Yelena Drexler, Alessia Fornoni*

*University of Michigan, Ann Arbor, MI, USA*: Jeffrey Hodgin^+^, Andrea Oliverio*

*University of Pennsylvania, Philadelphia, PA, USA*: Jon Hogan, Lawrence Holzman**, Matthew Palmer ^+^, Gaia Coppock

*University of Pittsburgh School of Medicine, Pittsburgh, PA, USA*: Michael Mortiz* *University of Washington, Seattle, WA, USA*: Charles Alpers^+^, J. Ashley Jefferson* *UT Southwestern, Dallas, TX, USA*: Kamal Sambandam*, Bethany Roehm

### Data Coordinating Center (DCC)

*Cedar Sinai Medical Center, Los Angeles, CA, USA*: Cynthia Nast^+^, Jean Hou^+^

*Duke University, Durham, NC, USA*: Laura Barisoni *Cleveland Clinic, Cleveland, OH, USA:* Crystal Gadegbeku** *Northwestern University, Chicago, IL, USA:* Abigail Smith**

*University of Michigan, Ann Arbor, MI, USA*: Brenda Gillespie, Bruce Robinson, Matthias Kretzler, Zubin Modi, Laura Mariani**

### Steering Committee Chair

Lisa M. Guay-Woodford, Children’s Hospital of Pennsylvania, Philadelphia, PA, USA

## Supplementary

**Figure 1.**
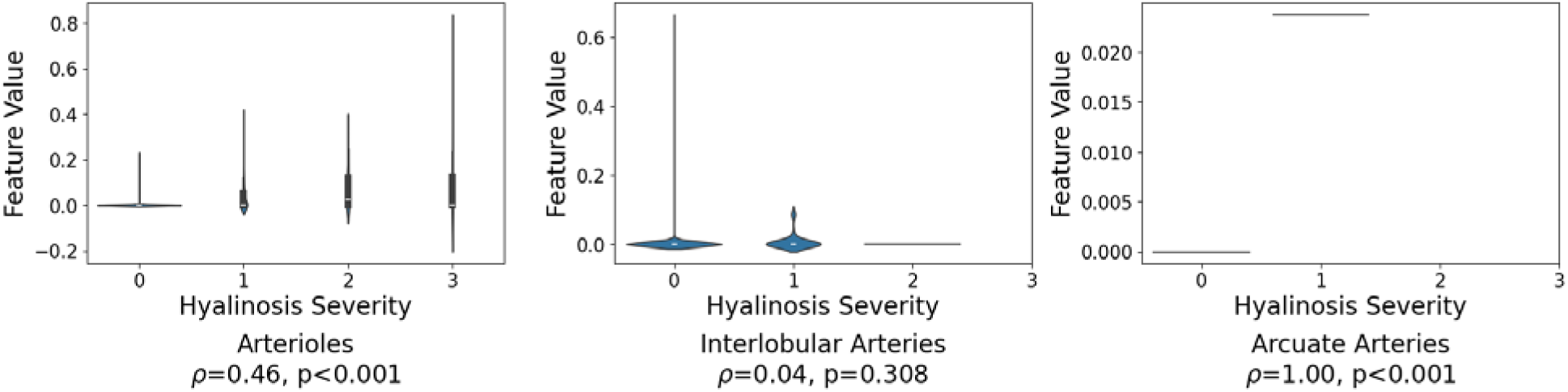
Violin plot of hyalinosis area ratio distribution across hyalinosis severity scores for arterioles (left), interlobular arteries (middle), and arcuate arteries (right).

**Figure 2.**
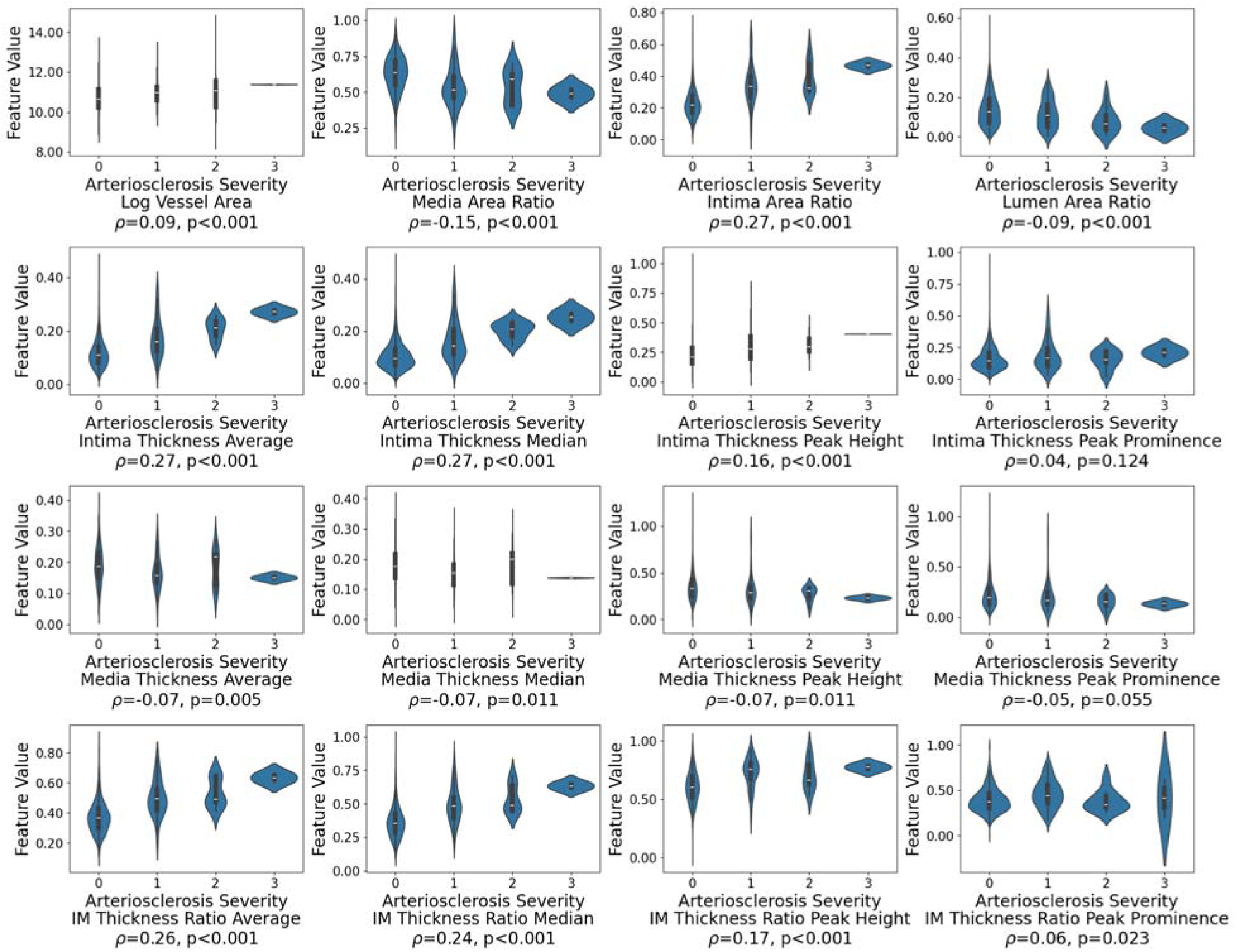
Violin plots of intra-arterial/arteriolar area and thickness-based features across arteriosclerosis severity scores for arterioles.

**Figure 3.**
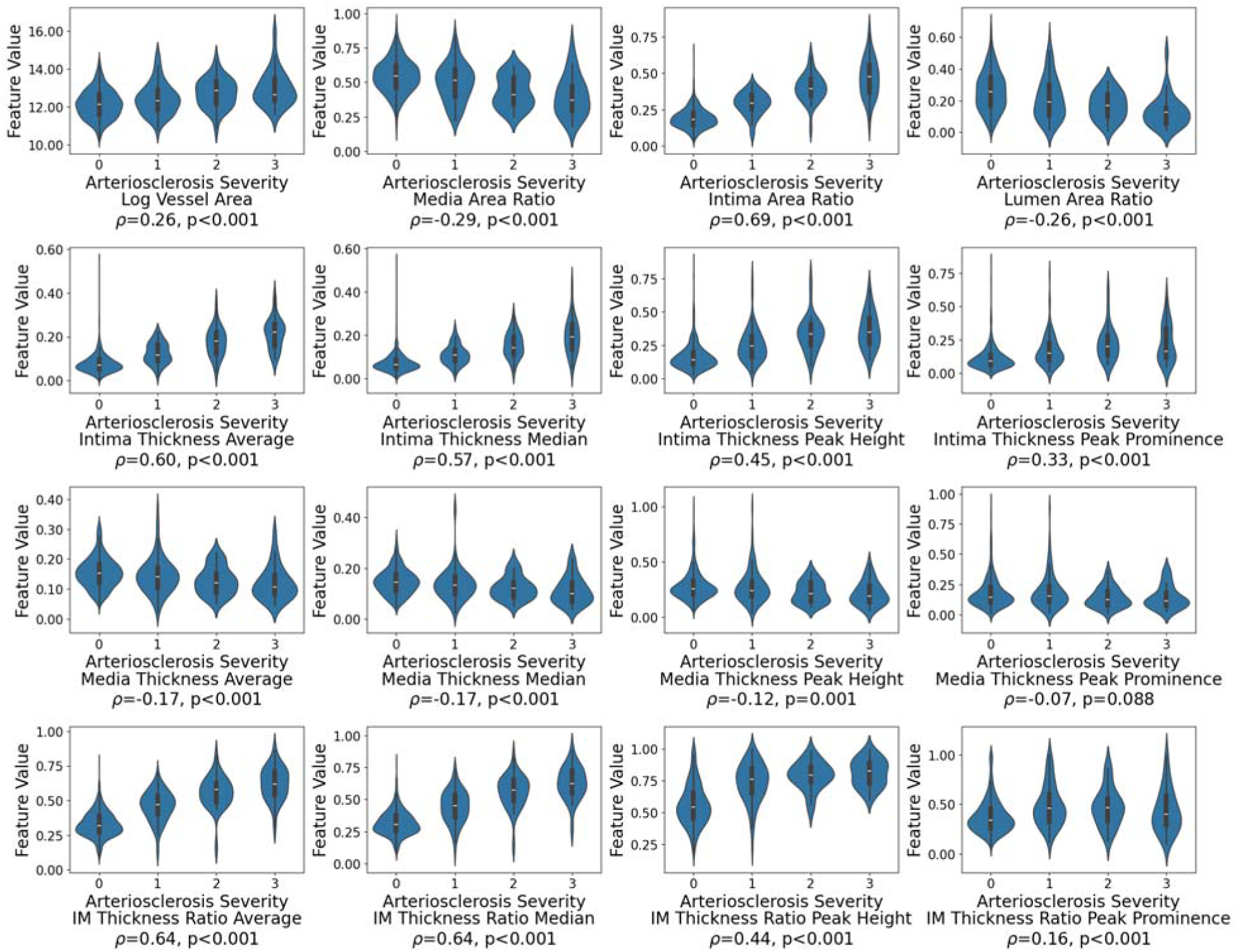
Violin plots of intra-arterial/arteriolar area and thickness-based features across arteriosclerosis severity scores for interlobular arteries.

**Figure 4.**
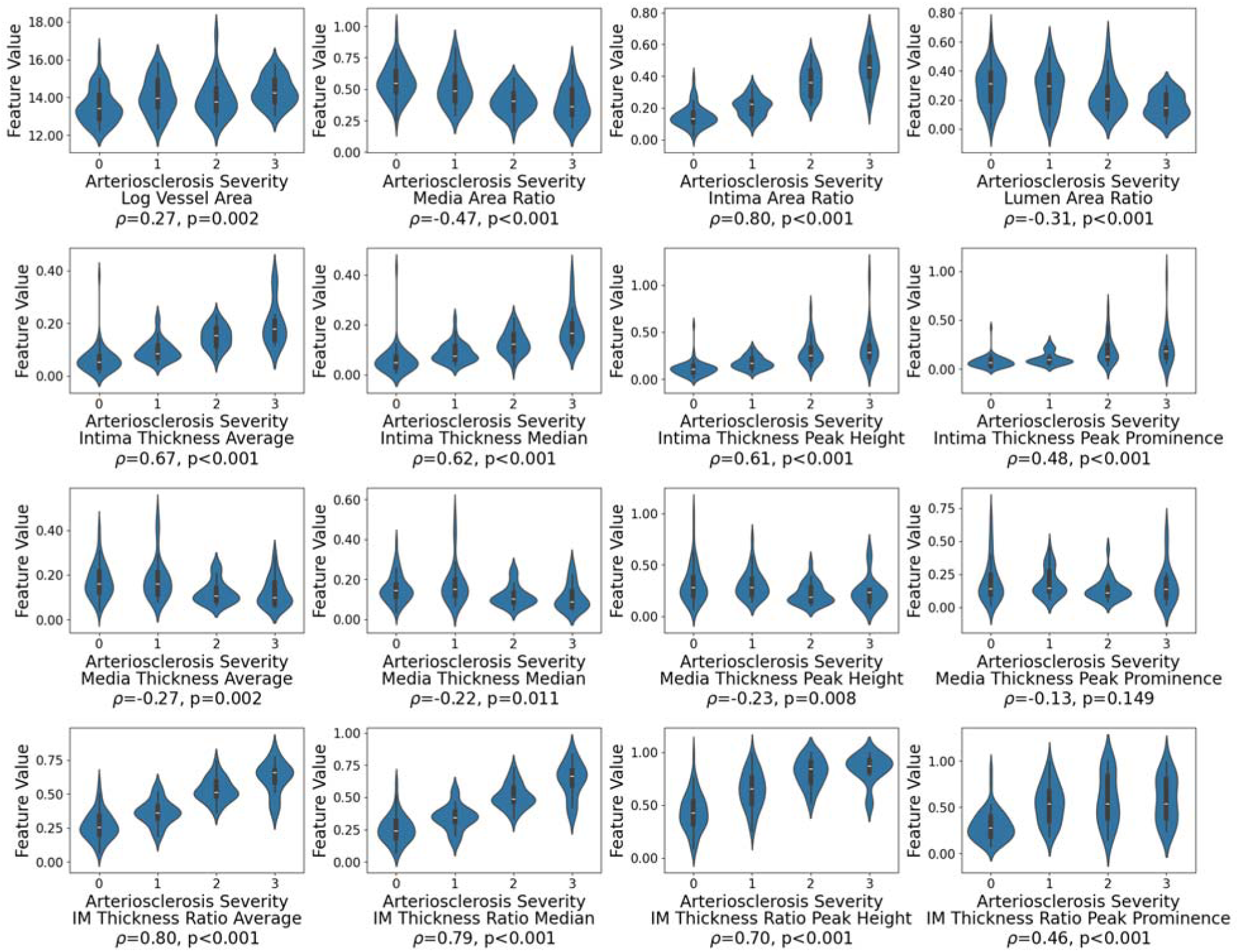
Violin plots of intra-arterial/arteriolar area and thickness-based features across arteriosclerosis severity scores for arcuate arteries.

## References

1. Baidoshvili, A. et al. Evaluating the benefits of digital pathology implementation: time savings in laboratory logistics. Histopathology 73, 784–794 (2018).

2. Barisoni, L. & Hodgin, J. B. Digital pathology in nephrology clinical trials, research, and pathology practice. Curr. Opin. Nephrol. Hypertens. 26, 450–459 (2017).

3. Barisoni, L., Lafata, K. J., Hewitt, S. M., Madabhushi, A. & Balis, U. G. J. Digital pathology and computational image analysis in nephropathology. Nat. Rev. Nephrol. 16, 669–685 (2020).

4. Brachtel, E. & Yagi, Y. Digital imaging in pathology--current applications and challenges. J. Biophotonics 5, 327–335 (2012).

5. Pantanowitz, L. et al. Twenty Years of Digital Pathology: An Overview of the Road Travelled, What is on the Horizon, and the Emergence of Vendor-Neutral Archives. J. Pathol. Inform. 9, 40 (2018).

6. Pantanowitz, L. et al. Validating whole slide imaging for diagnostic purposes in pathology: guideline from the College of American Pathologists Pathology and Laboratory Quality Center. Arch. Pathol. Lab. Med. 137, 1710–1722 (2013).

7. Retamero, J. A., Aneiros-Fernandez, J. & Del Moral, R. G. Complete Digital Pathology for Routine Histopathology Diagnosis in a Multicenter Hospital Network. Arch. Pathol. Lab. Med. 144, 221–228 (2020).

8. Tellez, D. et al. Quantifying the effects of data augmentation and stain color normalization in convolutional neural networks for computational pathology. Med. Image Anal. 58, 101544 (2019).

9. Aeffner, F. et al. Quantitative assessment of pancreatic cancer precursor lesions in IHC-stained tissue with a tissue image analysis platform. Lab. Invest. 96, 1327–1336 (2016).

10. Aeffner, F. et al. Validation of a Muscle-Specific Tissue Image Analysis Tool for Quantitative Assessment of Dystrophin Staining in Frozen Muscle Biopsies. Arch. Pathol. Lab. Med. 143, 197–205 (2018).

11. Blacher, S. et al. Quantitative Assessment of Mouse Mammary Gland Morphology Using Automated Digital Image Processing and TEB Detection. Endocrinology 157, 1709–1716 (2016).

12. Chen, J.-M. et al. Computer-aided prognosis on breast cancer with hematoxylin and eosin histopathology images: A review. Tumor Biol. 39, 1010428317694550 (2017).

13. Leo, P. et al. Computationally Derived Cribriform Area Index from Prostate Cancer Hematoxylin and Eosin Images Is Associated with Biochemical Recurrence Following Radical Prostatectomy and Is Most Prognostic in Gleason Grade Group 2. Eur. Urol. Focus 7, 722–732 (2021).

14. Li, X. et al. Deep learning segmentation of glomeruli on kidney donor frozen sections. J. Med. Imaging 8, 067501 (2021).

15. Wuttisarnwattana, P., Gargesha, M., van’t Hof, W., Cooke, K. R. & Wilson, D. L. Automatic Stem Cell Detection in Microscopic Whole Mouse Cryo-Imaging. IEEE Trans. Med. Imaging 35, 819–829 (2016).

16. Chen, Y. et al. Clinical Relevance of Computationally Derived Attributes of Peritubular Capillaries from Kidney Biopsies. Kidne*y360* 4, 648 (2023).

17. Fan, F. et al. Clinical Relevance of Computationally Derived Tubular Features: Spatial Relationships and the Development of Tubulointerstitial Scarring in MCD/FSGS. 2024.07.19.24310619 Preprint at 10.1101/2024.07.19.24310619 (2024).

18. Lucarelli, N. et al. Correlating Deep Learning-Based Automated Reference Kidney Histomorphometry with Patient Demographics and Creatinine. Kidne*y360* 4, 1726 (2023).

19. Hölscher, D. L. et al. Next-Generation Morphometry for pathomics-data mining in histopathology. Nat. Commun. 14, 470 (2023).

20. Border, S. et al. Improving quantification of renal fibrosis using deep-DUET. in Medical Imaging 2023: Digital and Computational Pathology vol. 12471 102–106 (SPIE, 2023).

21. Ginley, B. et al. Automated Computational Detection of Interstitial Fibrosis, Tubular Atrophy, and Glomerulosclerosis. J. Am. Soc. Nephrol. 32, 837 (2021).

22. Zhou, J. et al. Characterization of arteriosclerosis based on computer-aided measurements of intra-arterial thickness. J. Med. Imaging 11, 057501 (2024).

23. Aeffner, F. et al. Commentary: Roles for Pathologists in a High-throughput Image Analysis Team. Toxicol. Pathol. 44, 825–834 (2016).

24. Aeffner, F. et al. Introduction to Digital Image Analysis in Whole-slide Imaging: A White Paper from the Digital Pathology Association. J. Pathol. Inform. 10, 9 (2019).

25. Denic, A. et al. Prognostic Implications of a Morphometric Evaluation for Chronic Changes on All Diagnostic Native Kidney Biopsies. J. Am. Soc. Nephrol. 33, 1927 (2022).

26. Aeffner, F. et al. The Gold Standard Paradox in Digital Image Analysis: Manual Versus Automated Scoring as Ground Truth. Arch. Pathol. Lab. Med. 141, 1267–1275 (2017).

27. Adam, B. et al. Banff Initiative for Quality Assurance in Transplantation (BIFQUIT): Reproducibility of Polyomavirus Immunohistochemistry in Kidney Allografts. Am. J. Transplant. 14, 2137–2147 (2014).

28. Haas, M. et al. Banff 2013 Meeting Report: Inclusion of C4d-Negative Antibody-Mediated Rejection and Antibody-Associated Arterial Lesions. Am. J. Transplant. 14, 272–283 (2014).

29. Roberts, C. A. et al. Interpretive disparity among pathologists in breastsentinel lymph node evaluation. Am. J. Surg. 186, 324–329 (2003).

30. Roberts, J. M. et al. High reproducibility of histological diagnosis of human papillomavirus-related intraepithelial lesions of the anal canal. Pathology (Phila.) 47, 308–313 (2015).

31. Gadegbeku, C. A. et al. Design of the Nephrotic Syndrome Study Network (NEPTUNE) to evaluate primary glomerular nephropathy by a multidisciplinary approach. Kidney Int. 83, 749–756 (2013).

32. Zee, J. et al. Kidney Biopsy Features Most Predictive of Clinical Outcomes in the Spectrum of Minimal Change Disease and Focal Segmental Glomerulosclerosis. J. Am. Soc. Nephrol. 33, 1411 (2022).

33. Jayapandian, C. P. et al. Development and evaluation of deep learning–based segmentation of histologic structures in the kidney cortex with multiple histologic stains. Kidney Int. 99, 86–101 (2021).

34. Bankhead, P. et al. QuPath: Open source software for digital pathology image analysis. Sci. Rep. 7, 16878 (2017).

35. Labelbox | Data factory for the next GenAI. https://labelbox.com.

36. Naesens, M. et al. The Banff 2022 Kidney Meeting Report: Reappraisal of microvascular inflammation and the role of biopsy-based transcript diagnostics. Am. J. Transplant. 24, 338– 349 (2024).

37. Srivastava, A. et al. The Prognostic Value of Histopathologic Lesions in Native Kidney Biopsy Specimens: Results from the Boston Kidney Biopsy Cohort Study. J. Am. Soc. Nephrol. JASN 29, 2213–2224 (2018).

38. Eadon, M. T. et al. Kidney Histopathology and Prediction of Kidney Failure: A Retrospective Cohort Study. Am. J. Kidney Dis. Off. J. Natl. Kidney Found. 76, 350–360 (2020).

